# A comparison of alternative vaccination strategies for protecting those vulnerable to illness, hospitalization, or death upon infection with SARS-CoV-2

**DOI:** 10.1101/2025.08.30.25334767

**Authors:** Zhilan Feng, Troy Day, John W Glasser

## Abstract

U.S. vaccination policymakers initially prioritized healthcare, other frontline essential workers, and those at greatest risk of severe COVID-19. Protection may be direct, by decreasing susceptibility to infection if exposed, or indirect, by decreasing the risk of exposure through the vaccination of oconsistent with the literature, and improvedthers. We modified our published SARS-CoV-2 transmission model by including immunity-moderated disease, updating some parameters, and making others time-varying. Then we derived the metapopulation reproduction numbers and related analytical quantities and compared the respective impacts of protecting vulnerable people directly and indirectly from 14 December 2020, when vaccination began, to 11 May 2024, when the U.S. Public Health Emergency officially ended. We found that proportionately more young people, particularly adolescents, had antibodies to nucleocapsid protein than adults before schools reopened for the 2021-22 school year, their vaccination lagged far behind that of older people, especially during the Omicron epoch, and they caused the most secondary infections *per capita*. Nonetheless, while vaccinating younger people would have reduced transmission more, we found that – if the effectiveness of vaccination and duration of vaccine-induced immunity were age-independent, as limited available evidence suggests – the actual strategy averted more illnesses, hospitalizations, and deaths. Because of the age-dependence of severe COVID-19, which policy was designed to mitigate, evidently vaccinating older people was more effective than vaccinating the younger ones who might otherwise have infected them. This result would not hold if, due to immune senescence, the effectiveness of vaccination or duration of vaccine-induced immunity varied inversely with age.

**Highlights:**

- published SARS-CoV-2 transmission model updated for the entire COVID-19 pandemic
- derived NGMs whose dominant eigenvalues are the **ℛ**_**0**_ **and ℛ**_***E***_ associated quantities
- motivated by **∇ ℛ**_***E***_, compared the actual vaccination program, designed to mitigate disease, with one designed to mitigate transmission
- formulated alternative program in proportion to the left eigenvector associated with **ℛ**_***E***_ versus **−∇ ℛ**_***E***_, eliminating the need to assume proportional mixing
- assuming that the efficacy and duration of protection were age-independent, as limited available evidence suggests, the actual vaccination program was more effective
- nonetheless, vaccinating to reduce transmission would have other benefits

## I. Introduction

During respiratory disease pandemics, healthcare and other essential workers are typically at the greatest risk of infection. In contrast, older adults and immunocompromised individuals face the greatest risk of severe outcomes, such as hospitalization and death, if infected. While prior exposure to related influenza A viruses provided many adults with partial immunity during the H1N1 pandemic in 2009 (Krammer 2019), such baseline immunity was largely absent in the population with few people having antibodies to SARS-CoV-2 prior to the COVID-19 pandemic (Althoff, et al. 2022).

In response, U.S. vaccine policymakers recommended prioritizing healthcare workers, other frontline essential workers, and individuals at the greatest risk of severe disease when COVID-19 vaccines became available in late 2020 and early 2021 (Dooling, et al. 2020). Vaccine strategies were designed to consider only direct protection―by reducing susceptibility to infection if exposed or severity of disease if infected—not indirect protection―by reducing the risk of exposure through the vaccination of others.

Because the prevalence of co-morbidities that increase the risk of symptoms varies with age (Clark, et al. 2020), infections of younger individuals, who generally experienced mild symptoms or remained asymptomatic, were under-reported (Couture, et al. 2022). However, person-to-person contact rates amongst school-aged children and adolescents are substantially higher than those of other age groups (Mossong, et al. 2008). While most person-to-person contacts occur among people roughly the same age, inter-generational (i.e., parents with children and grandparents with grandchildren) and age-independent workplace contacts (Glasser, et al. 2012; Feng, et al. 2017) also occur non-randomly (i.e., mixing is preferential, table 1). Thus, older adults who are at risk of severe disease may contact younger individuals at home or in the workplace who are infectious but asymptomatic.

**Table 1.** Glossary.

| Term | Symbol | Definition |
| --- | --- | --- |
| Marginal contact rates | $a_i$ | *Per capita* numbers of contacts per day (or another period) regardless of with whom |
| Proportionate (or proportional) mixing | $a_i N_i / \sum_j a_j N_j$ | Random (i.e., the probability of contacting a member of group $i$ is the quotient of their contacts, *per capita* rate $a \times$ number of people $N$, and that of all groups) |
| Preferential mixing |  | Nonrandom (e.g., assortative, disassortative) |
| Forces of infection | $\lambda_i(t) = a_i \beta_i \sum_j c_{ij} \frac{I_j(t)}{N_j}$ | Rates at which susceptible people in groups $i$ are infected, a function of their contact rates $a_i$, probabilities of infection upon contact with infectious people $\beta_i$, proportions of their contacts that are with members of the $j^{\text{th}}$ group $c_{ij}$, and that group's attack rate (i.e., proportion infectious, $I_j(t)/N_j$) |
| Gradient | $\nabla$ | The gradient of a multi-variable function is the vector of partial derivatives with respect to the variables |
| Metapopulation |  | A population composed of sub-populations (e.g., age groups in each of several locations) |
| Metapopulation reproduction numbers | $\mathcal{R}_0$ and $\mathcal{R}_E$ | Potential, $\mathcal{R}_0$ and actual, $\mathcal{R}_E$ average numbers of secondary infections per primary |
| Next-generation matrix | NGM | A matrix (e.g., Leslie matrix in population biology) whose largest eigenvalue is $\mathcal{R}_0$ or $\mathcal{R}_E$ |
| Sub-population contributions to $\mathcal{R}_0$ or $\mathcal{R}_E$ | | The left eigenvector associated with the largest eigenvalue of the NGM |

First we introduce our latest model of SARS-CoV-2 transmission and control in the United States. Then we describe analytical results that motivated us to wonder if those most vulnerable to serious illness, hospitalization, and death might have been better protected indirectly than directly, by the vaccination of those who might otherwise infect them than by vaccination themselves. Accordingly, by simulating our model during the entire Public Health Emergency, we compared the implemented vaccination strategy’s impact with that of a hypothetical alternative designed to reduce exposures and thereby to protect those most vulnerable to serious disease indirectly. We conclude by placing results in context.

## II. Methods

As the biology of COVID-19 became increasingly well understood, we modified successive age- and location-stratified Susceptible-Exposed-Infectious-Recovered (SEIR) models to include newly appreciated features that might affect transmission of SARS-CoV-2 among members of the U.S. population if we could model them accurately. We have reduced models only to facilitate computation.

Our current model describes later pandemic epochs, during which those partially immune may have experienced milder disease upon infection than immunologically naïve people, better than our penultimate model (Glasser, et al. 2023; Vo, et al. 2023). Time-varying parameters describe host behavior, availability of less effective vaccines, and pathogen evolution (Supplement).

### II.1 Model

The model diagrammed in figure 1 can be represented as a system of ordinary differential equations (ODEs), where *S*_*i*_ is the number who are susceptible in group *i* (here *i* or *j* index ages, locations, or combinations; there are *n* such groups); *E*_*is*_ and *E*_*ip*_ are the numbers immunologically naïve and partially immune who have been infected, but are not yet infectious; *V*_*i*_ is the number who have received one dose of vaccine (≥14 days ago); *I*_*iw*_ (*w = a, pm, m, ps, s, h*) are the numbers whose infections are mild, pre-moderate, moderate, pre-severe, severe, or who are hospitalized; *R*_*i*_ is the number who have recovered or been vaccinated a second time (≥14 days ago), and are temporarily immune; *P*_*i*_ is the number whose immunity has waned, so can be (re)infected or have their immunity boosted by (re)vaccination; and *M*_*i*_ is the number who have died:

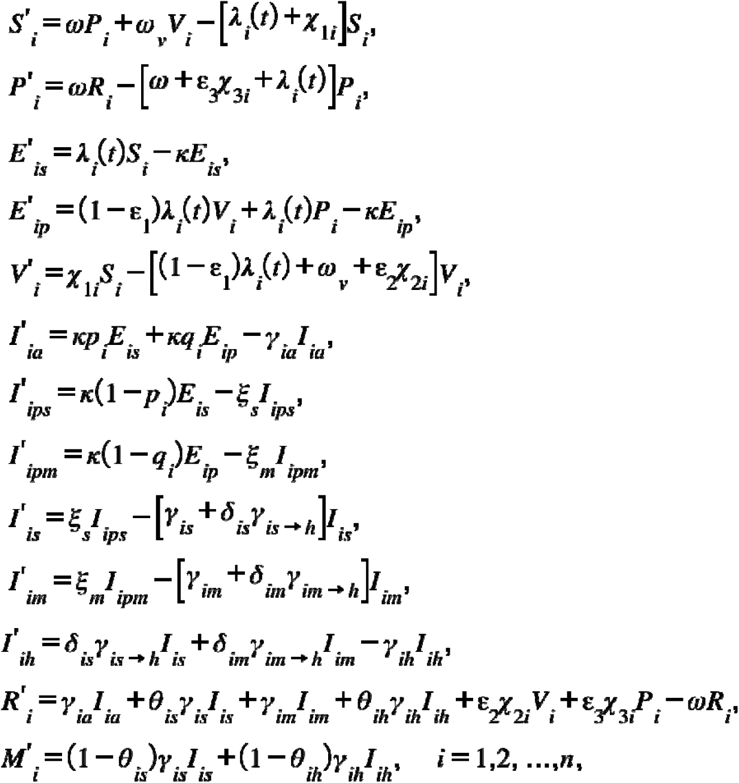

where the force or hazard rate of infection among susceptible people in group *i*, those having recently received one vaccine dose (whose susceptibility to infection is 1 – □_1_, where □_1_ is the effectiveness of dose 1) or who were infected or fully vaccinated some time ago, but whose immunity has waned, is 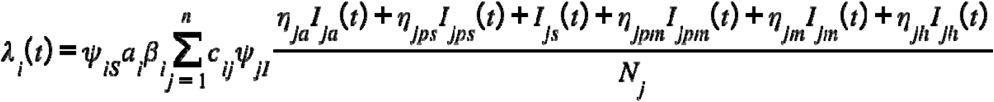, where ***N***_***j***_**=*S***_***j***_ **+*P***_***j***_ **+*E***_***js***_ **+*E***_***jp***_ **+*V***_***j***_ **+*I***_***ja***_ **+*I***_***jps***_ **+*I***_***jpm***_ **+*I***_***js***_ **+*I***_***jm***_ **+*I***_***jh***_ **+*R***_***j***_ **+*M***_***j***_, and ***i***,***j*=1**,**2**,**…**,***n***.

**Figure 1.**
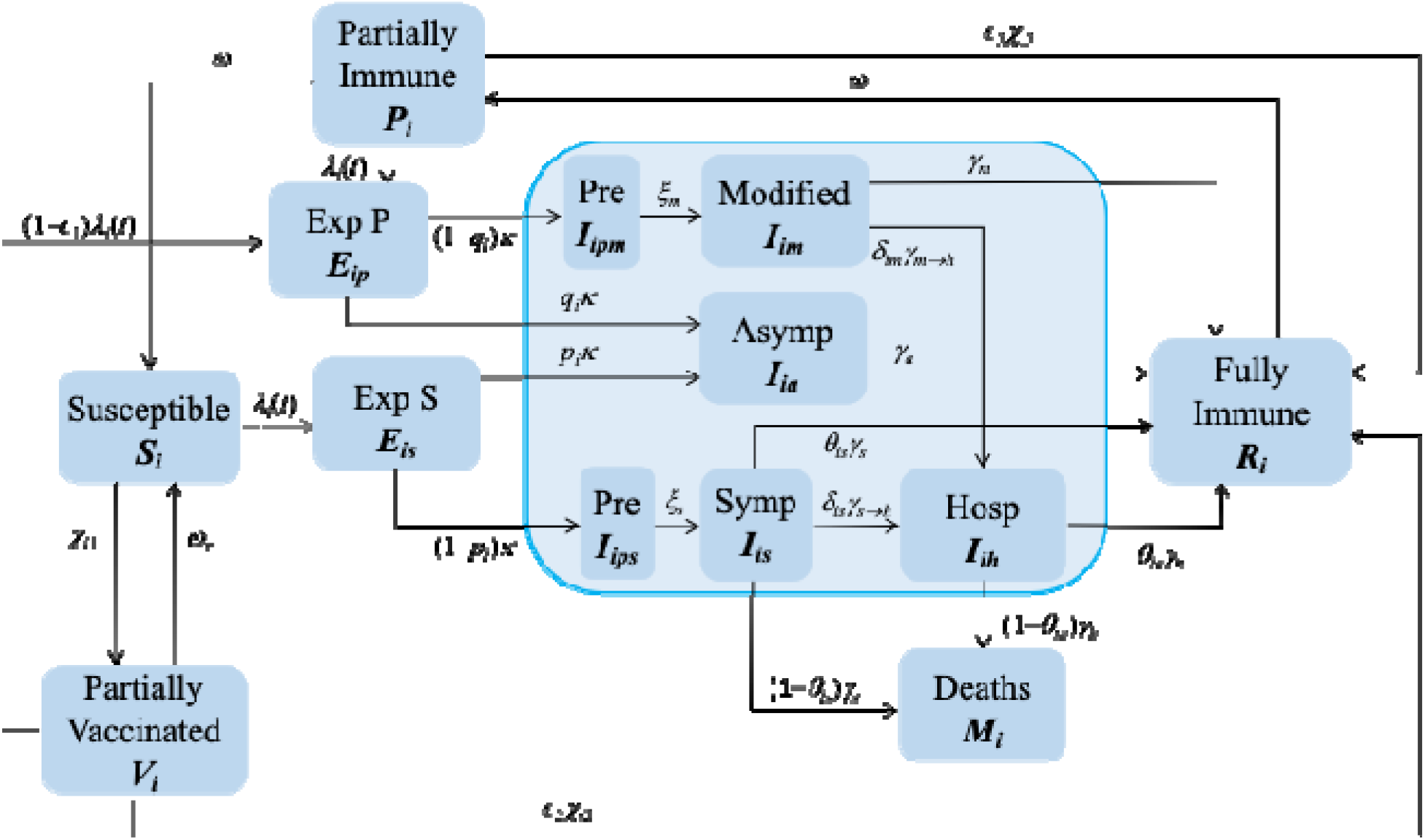
A Susceptible-Exposed-Infectious-Recovered (SEIR) model with mild, moderate, and severe disease. While the model described by Glasser, et al. (2023) and Vo, et al. (2023) was suitable early in the pandemic, immunity-moderated disease soon became a feature of COVID-19.

In this expression, ***ψ***_***iS***_ **= 1−*f***_***i***_ ***b***_***iS***_ and ***ψ***_***jI***_ **= 1−*f***_***j***_ ***b***_***jI***_ represent reductions in susceptibility and infectiousness, respectively, due to physical-distancing if possible and mask-wearing otherwise; *f*_*i*_ is the proportion of group *i* that comply with such recommendations, and *b*_*iS*_ and *b*_*iI*_ are their respective efficacies. Here *a*_*i*_ is the *per capita* contact rate; *β*_*i*_ is the probability of transmission per contact between susceptible and infectious people (*a*_*i*_ × *β*_*i*_ is the effective contact rate); *c*_*ij*_ is the proportion of their contacts that members of group *i* have with those of group *j*; and *N*_*j*_ is the number in that group. As *M*_*j*_ is small relative to *N*_*j*_, we maintain 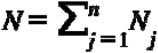 constant by including *M*_*j*_ in the equation for *N*_*j*_.

### II.2 Parameters

The Centers for Disease Control and Prevention (CDC) and Advisory Committee on Immunization Practices (ACIP) initially recommended prioritizing COVID-19 vaccination of health care personnel and residents of long-term care facilities, then people aged ≥ 75 years and other frontline essential workers, people aged 65-74 years, ones aged 16-64 years with high-risk medical conditions, and essential workers not included earlier (Dooling, et al. 2021).

While state and local health departments considered these recommendations, some modified the groups prioritized, or how quickly phase transitions occurred. As most people in prioritized groups were vaccinated and supply increased, CDC/ACIP recommendations were expanded to include others aged ≥ 16 years (Oliver, et al. 2023; Dooling, et al. 2021). Rather than programming these details, we calculated age-, location-, and dose-specific vaccination rates from the weekly numbers of vaccinations by age and dose that each state and the District of Columbia (D.C.) reported to the CDC.

As several investigators had reported reduced vaccine effectiveness with age after the first dose (e.g., Collier, et al. 2021; Rosenberg, et al. 2022), but not after the second (Jergović, et al. 2022) or third (Newman, et al. 2022; Renia, et al., 2022), we assumed that vaccine effectiveness and the duration of immunity (infection-as well as vaccination-induced) were age independent.

We analyzed information from two nationwide, serial, cross-sectional serological surveys using stable assays, one of antibodies to nucleocapsid protein (anti-N) among commercial laboratory clients (Bajema, et al. 2021) and other of antibodies to spike protein (anti-S) as well among blood-donors (Jones, et al. 2021). We a) calculated the forces of infection (table 1), attack rates (table 1), and – together with information about interpersonal contacts in the United States from Prem, et al. (2017) – probabilities of infection upon contact (table 1) and b) initialized our penultimate model from the first serosurvey during Q4 of 2020 (Glasser, et al. 2023).

All other parameters except the effectiveness of NPIs at reducing susceptibility if susceptible, and infectiousness if infected, were informed by the literature (definitions in table 2, values and references in the Supplement). We calibrated our penultimate model by setting those unknown-if-knowable parameters equal to each other and choosing a value with which predicted infections resembled the prevalence of anti-N just before vaccination began. Predictions of our current model (figure 1), which includes immunity-moderated disease and differs in other ways (Supplement Section 2), subsequently converge on the prevalence of anti-S with or without anti-N from the second serosurvey (figure 2).

**Table 2.**
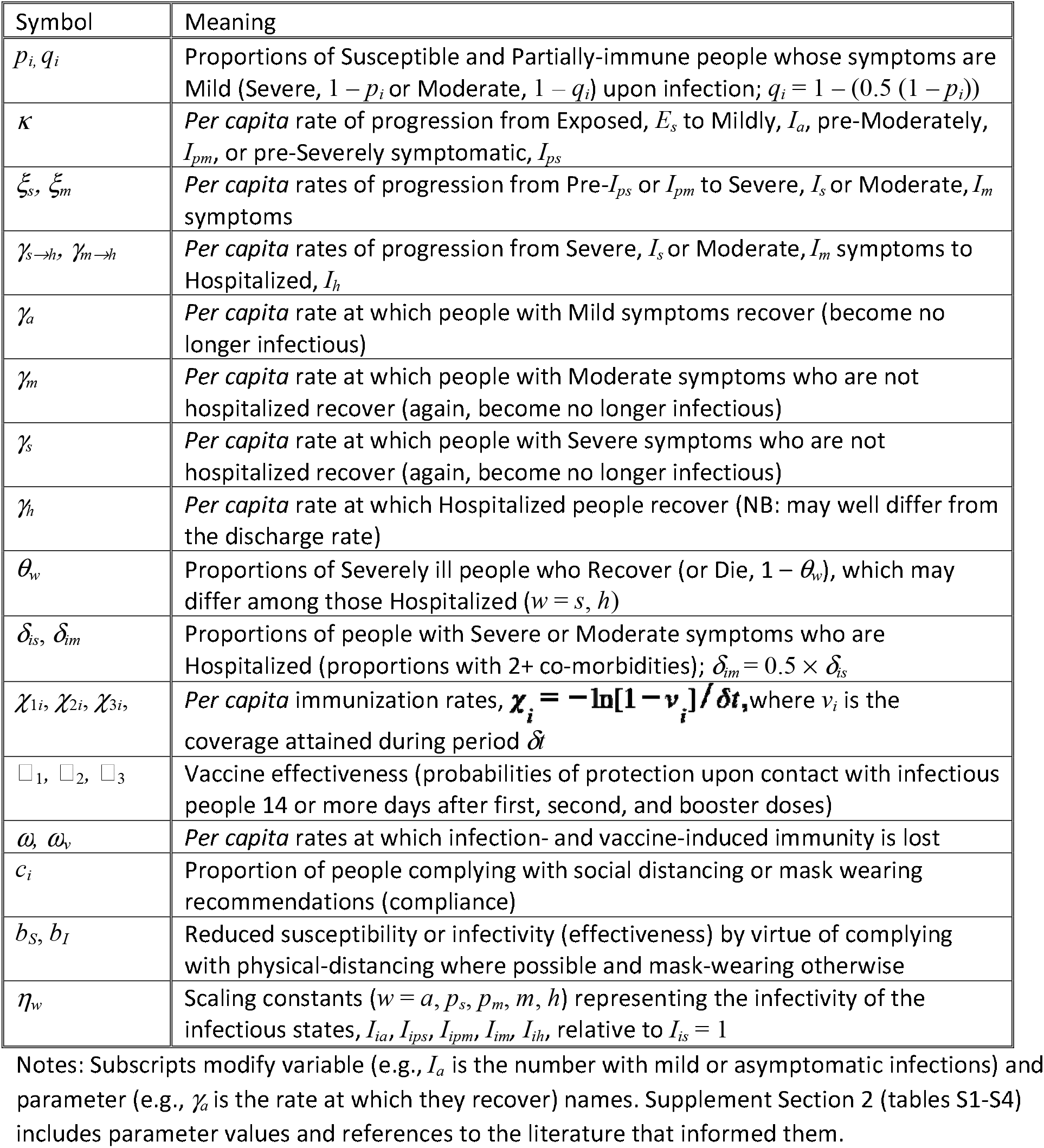
Parameter definitions.

**Figure 2.**
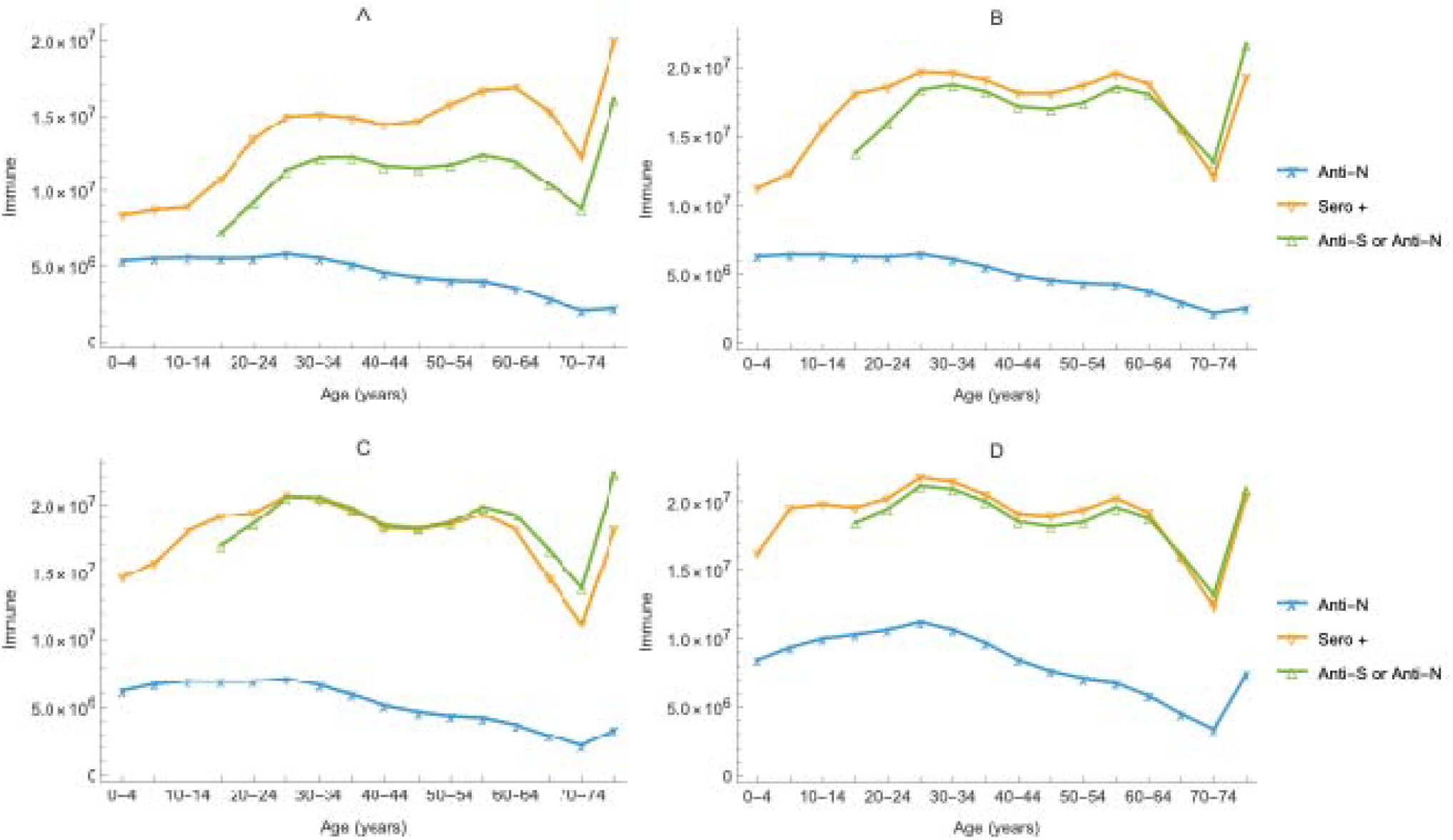
Comparison of predicted seropositivity (yellow curves) during the 1^st^, 2^nd^, 3^rd^, and 4^th^ quarters of 2021 (A, B, C, and D, respectively) and that observed (green curves) among blood donors (i.e., anti-N with or without anti-S). The blue line is anti-N among commercial laboratory clients. The oldest age class is an open interval, those aged 75+ years, which is larger than the other classes, whose width is 5 years.

### II.3 Analytical and Simulation Methods

In metapopulation (table 1) models, reproduction numbers (table 1) are properties of next-generation matrices, NGMs (table 1), which we derive in the usual way (Appendix). Their dominant eigenvalues are potential or actual average numbers of secondary infections per primary (table 1), denoted **ℛ**_**0**_ or **ℛ**_***E***_, respectively. Their associated left eigenvectors (table 1) are subpopulation contributions (Hurford, et al., 2010). In our models, effective reproduction numbers, **ℛ**_***E***_, differ from basic ones, **ℛ**_**0**_, insofar as NPIs, vaccination, and hospital infection-control procedures reduce the average number of secondary infections per primary.

The negative gradient of the effective reproduction number, **−∇ ℛ**_*E*_, with respect to control parameters (e.g., vaccination rates) indicates how to reduce **ℛ**_*E*_ most expeditiously (Feng, et al. 2015). Vo, et al. (2023) compared the respective impacts of three vaccination strategies during Q1 of 2021: actual, random, and rates that would minimize the metapopulation **ℛ**_*E*_ given available vaccine (Feng, et al. 2017, 2020). As calculating **−∇ ℛ**_*E*_ with that constraint proved difficult with their full model, they reduced its age structure from 16 to 6 classes. And, to obtain an expression for the metapopulation **ℛ**_*E*_ whose multivariate partial derivative could be calculated, they assumed proportional mixing (table 1), whereupon **ℛ**_*E*_ is the trace of the NGM (i.e., sum of its diagonal elements).

Motivated partly by the belief that realistic mixing is preferential (table 1) and partly by the fact that reduced models lack the dynamics of full ones, we compared the respective impacts of a) the reported weekly vaccinations by age and dose at each of 51 locations, the U.S. states and D.C., from 14 December 2020, when vaccination began in the United States, through the end of the Public Health Emergency on 11 May 2023, after which reports to the CDC became less reliable (Silk, et al. 2023) and b) reallocations by dose at each location among all 16 age classes, 0-4, 5-9, …, and 75+ years, in proportion to their contributions to the metapopulation **ℛ**_*E*_ the previous week.

To assess the impact of vaccination, we subtracted predicted illnesses, hospitalizations, and deaths among persons with vaccination from those without. To simulate the actual strategy, we converted weekly quotients of reported vaccinations by age, location, and dose, and those eligible from the model solution at the end of the previous week into rates (table 2), vaccinated daily at one seventh of those rates for a week, and repeated. To simulate the alternative strategy, we summed reported vaccinations by dose over all ages at each location, allocated them, again by dose, to age groups in proportion to the left eigenvector associated with the largest eigenvalue of the NGM at the end of the previous week, vaccinated daily at one seventh of the corresponding rates for a week, and repeated.

## III. Results

### III.1 Analytical Results

In their evaluation of the U.S. vaccination program, Vo, et al. (2023) were motivated by the declining gradient of the effective reproduction number with respect to vaccination rates among adults during January of 2021 (their figure 8). Nonetheless, they concluded that the actual program accomplished early policy goals better than two alternatives (their figure 9).

Our figures 3 compare **−∇ ℛ**_*E*_ before vaccination began and at the end of Q1 of 2021 (panels a and b), when it was positive overall and only among children and adolescents, respectively. These results suggest that another strategy might have proved superior later, especially if vaccination did not protect people at risk of severe disease as well or for as long as it protected others. Under those conditions, vaccinating people who otherwise could infect vulnerable ones might be more effective.

**Figure 3.**
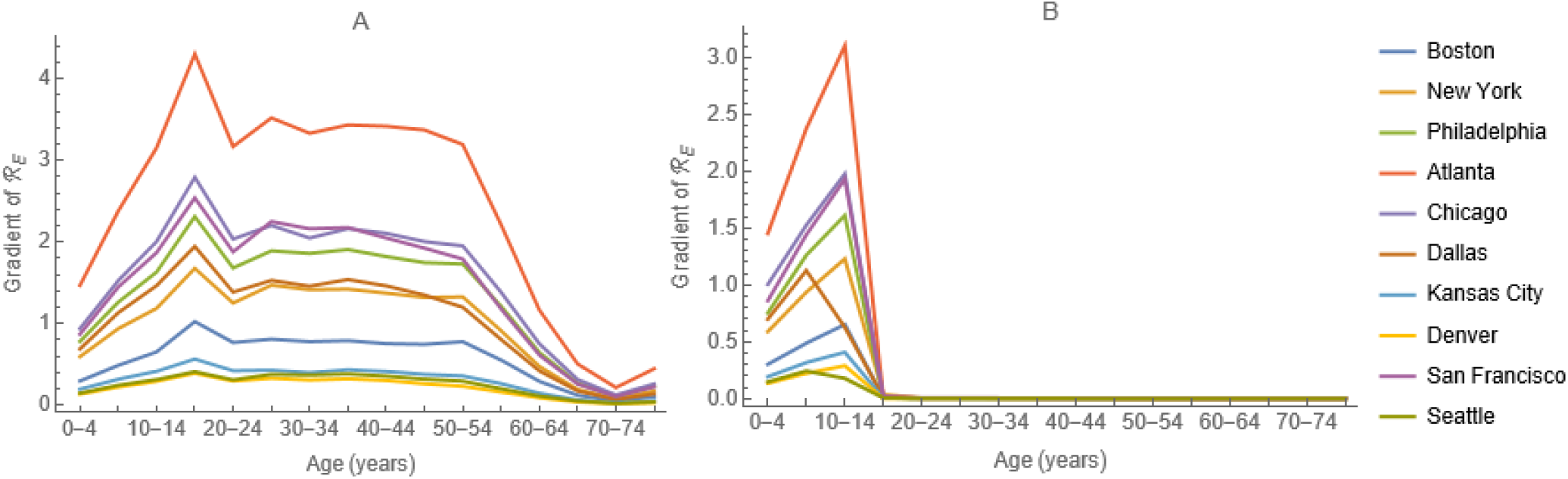
Gradient of the metapopulation effective reproduction number, **∇ ℛ**_***E***_ A) pre-vaccination and B) on 4 April 2021 in a model stratified by HHS Regions, denoted by headquarter locations.

Moreover, the age-profile of anti-N prior to schools reopening for the 2021-22 school year (figure 4b) mirrors that of the marginal contact rates (figure 4a), indicating that, while NPIs may have reduced contacts overall, those of school-aged children or adolescents were not reduced disproportionately. The peak indicates that adolescents were infected (and presumably infected others) with SARS-CoV-2 at the greatest rate (i.e., relative to younger and older people, were super-spreaders, Teicher 2023) while the shoulder (during the reproductive ages) indicates that children infected their parents or vice versa.

**Figure 4.**
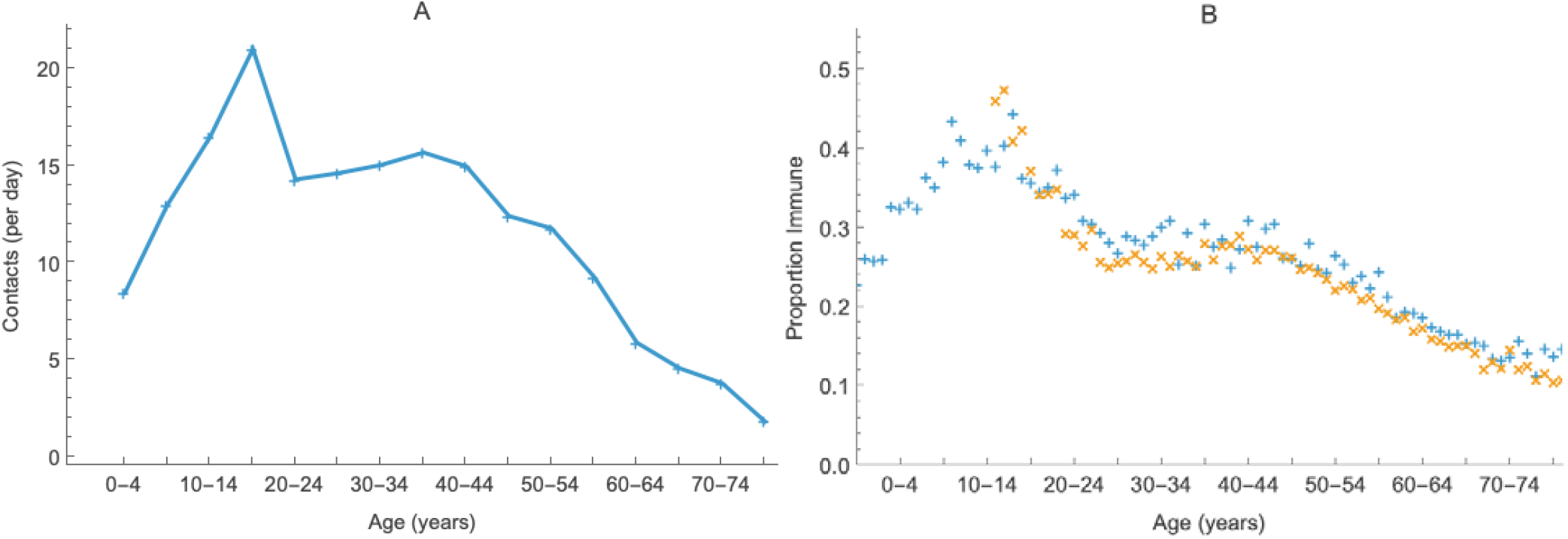
Comparison of the A) marginal contact rates (table 1) from Prem, et al. (2017) and B) proportions of commercial laboratory clients (Bajema, et al. 2021) and blood-donors (Jones, et al. 2021), the ‘+’ and ‘x’ symbols respectively, with antibodies to nucleocapsid protein during September of 2021.

Consequently, although viral load varies with symptoms (Marks, et al. 2021), analytical results suggest that vaccinating super-spreaders might have attained policy objectives more effectively than vaccinating those most vulnerable to serious illness, hospitalization, and death, especially if the immunological response to vaccination declines with age (Wang, et al. 2021).

### III.2 Simulation Results

Immunizations by dose and their respective impacts are illustrated during 2020-21 in figures 5 and 2022-23 in figures 6. Panels on the left (a and c) compare immunizations while panels on the right (b and d) compare disease outcomes (i.e., illnesses, hospitalizations, and deaths). We compare vaccination strategies (i.e., subtract alternative immunizations and disease events from actual ones) on figures 7 and **ℛ**_*E*_ for different strategies on figures 8. The left-side panels (a and c) compare weekly **ℛ**_*E*_ for the two strategies to each other and the threshold value of one while those on the right (b and d) compare proportionate reductions, (**ℛ**_*E*_ actual strategy –**ℛ**_*E*_ alternative strategy)/ **ℛ**_*E*_ actual strategy.

**Figures 5.**
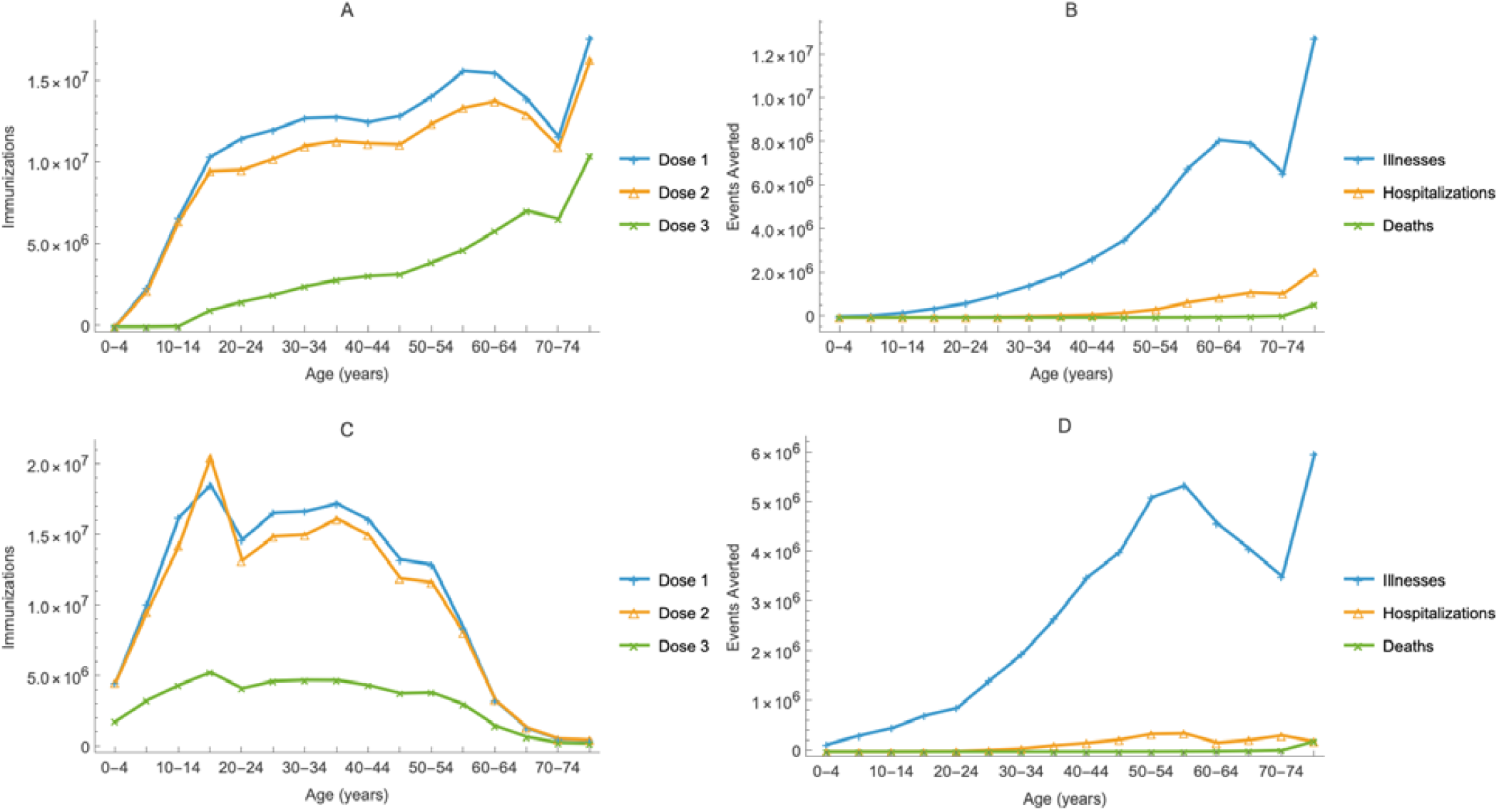
Immunizations by age and dose (A and C) and moderate plus severe illnesses, hospitalizations, and deaths averted by age (B and D) during 2020-21 by the actual (A and B) and alternative (C and D) vaccination strategies. We estimate that approximately 6×10^7^ illnesses, 6.8×10^6^ hospitalizations, and 7.3×10^5^ deaths were averted by the actual and 4.5×10^7^ illnesses, 2.3×10^6^ hospitalizations, and 2.7×10^5^ deaths by the alternative strategy.

**Figures 6.**
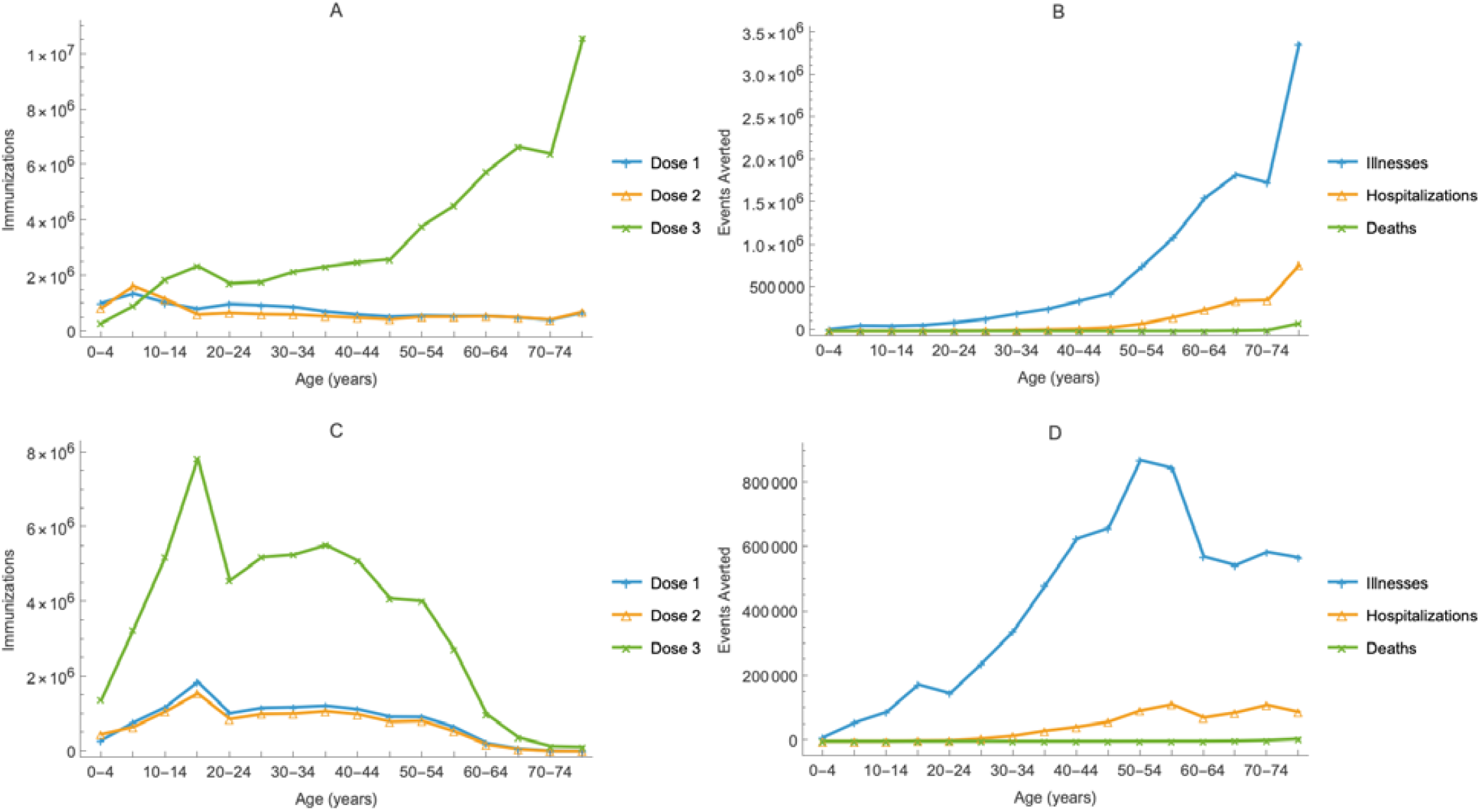
Immunizations by age and dose (A and C) and moderate plus severe illnesses, hospitalizations, and deaths averted by age (B and D) during 2022-23 by the actual (A and B) and alternative (C and D) strategies. We estimate that approximately 1.2×10^7^ illnesses, 2.1×10^6^ hospitalizations, and 1.1×10^5^ deaths were averted by the actual and 6.9×10^6^ illnesses, 7.4×10^5^ hospitalizations, and 1.2×10^4^ deaths by the alternative strategy.

**Figures 7.**
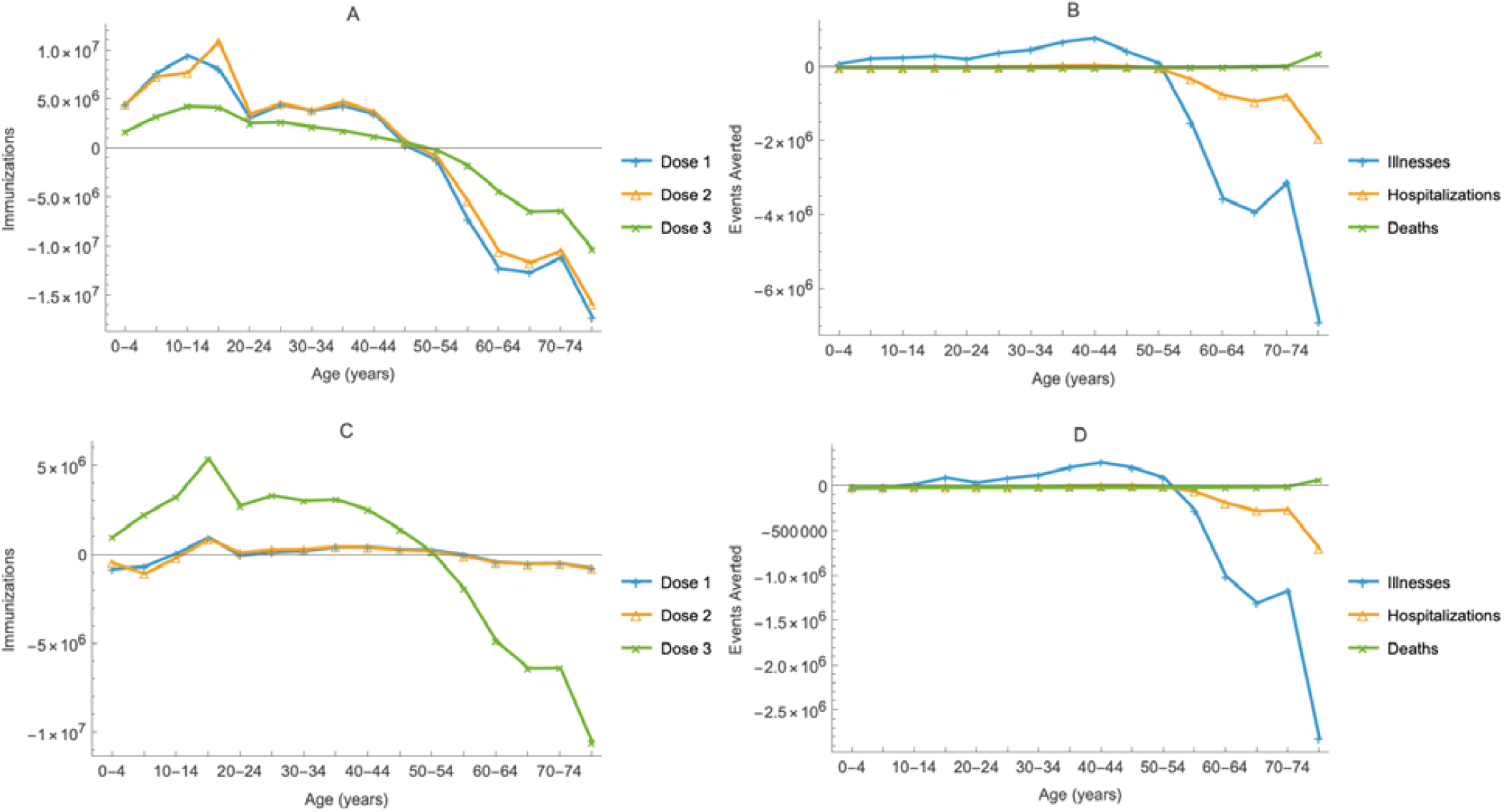
Difference between alternative and actual strategies in immunizations by age and dose (A and C) and moderate plus severe illnesses, hospitalizations, and deaths averted by age (B and D) during 2020-21 (A and B) and 2022-23 (C and D). Positive or negative values indicate in which ages and by how much the alternative immunizations by dose (A and C) or illnesses, hospitalizations, and deaths averted (B and D) are more or less than the actual strategy.

**Figures 8.**
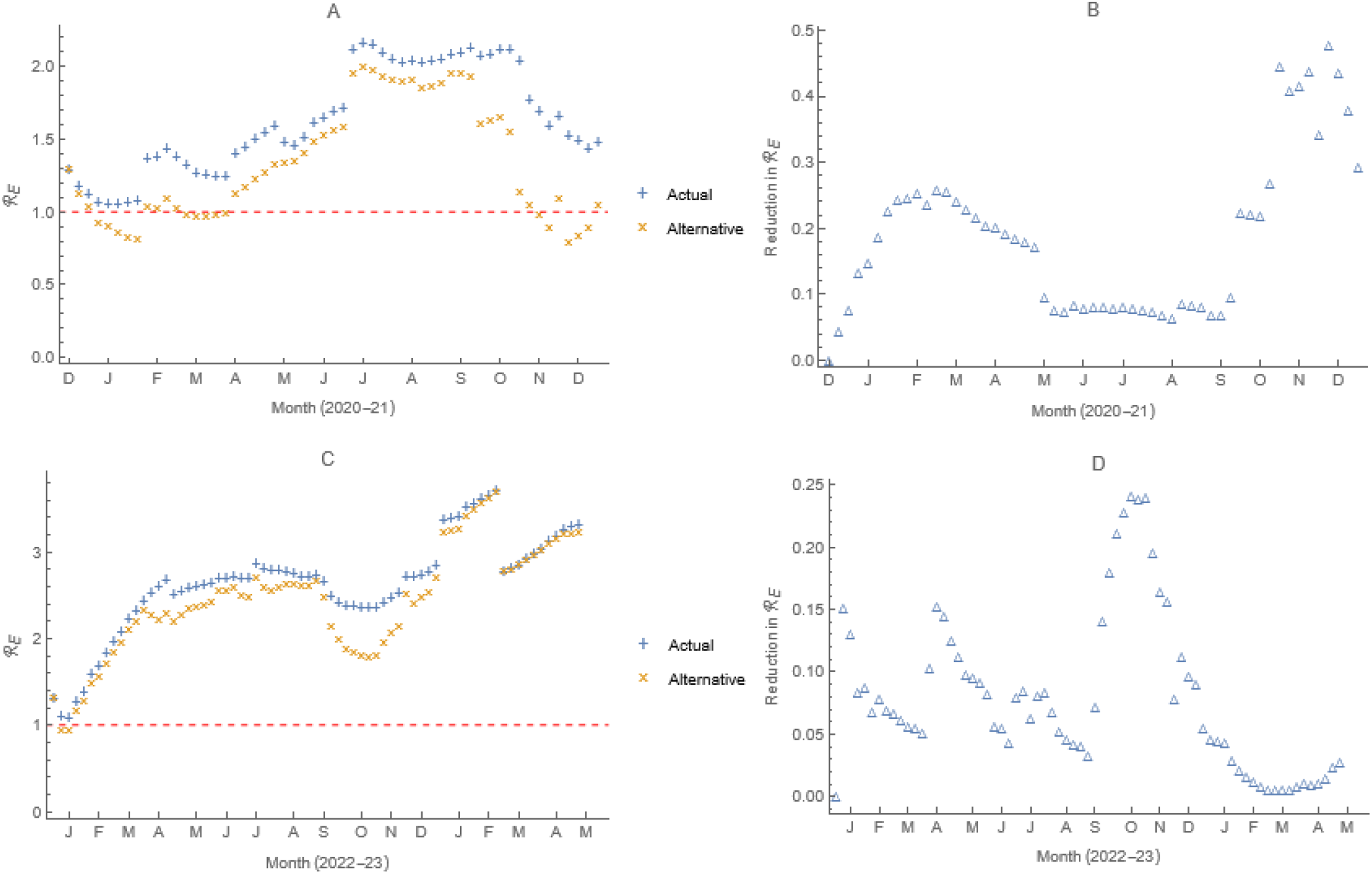
Weekly ***ℛ***_***E***_ for the two strategies (A and C) and proportionate reductions, **(*ℛ***_***E***_ **actual strategy −*ℛ***_***E***_ **alternative strategy)/ *ℛ***_***E***_ **actual strategy** (B and D) during 2020-21 (A and B) and 2022-23 (C and D). The red dashed horizontal line on figures A and C indicates ***ℛ***_***E***_ **=1**,the threshold above and below which new infections increase and decrease, respectively.

Both strategies reduced illnesses, hospitalizations, and deaths (figures 5 and 6, panels b and d). During 2020-21, we estimate that approximately 6×10^7^ illnesses, 6.8×10^6^ hospitalizations, and 7.3×10^5^ deaths were averted by the actual strategy and 4.5×10^7^ illnesses, 2.3×10^6^ hospitalizations, and 2.7×10^5^ deaths by the alternative strategy. During 2022-23, the corresponding numbers are lower, 1.2×10^7^ illnesses, 2.1×10^6^ hospitalizations, and 1.1×10^5^ deaths by the actual and 6.9×10^6^ illnesses, 7.4×10^5^ hospitalizations, and 1.2×10^4^ deaths by the alternative strategy, because there were fewer vaccinations during the later period and a greater proportion were booster versus primary doses (figures 5a and 6a).

Thus, as illustrated in figures 7b and d, we estimate that approximately 1.5×10^7^ more illnesses and 4.5×10^6^ more hospitalizations and 4.6×10^5^ more deaths were averted by the actual than alternative strategy during 2020-21. The corresponding numbers during 2022-23 are again lower, 5.2×10^6^ illnesses, 1.3×10^6^ hospitalizations, and 9.5×10^4^ deaths, for the same reasons.

Middle-aged people were vaccinated in both strategies (figures 5 and 6, panels a and c), with more younger individuals (aged < 50 years) being immunized in the alternative and older individuals (aged ≥ 50 years) in the actual strategy (figures 7a and c). Because the risk of the aforementioned disease outcomes for younger people is not as high as for their older counterparts (table S2), the alternative strategy’s impact is notable (figures 7b and d). More of the events averted must have been attributable to the vaccination of those who might otherwise have caused them (i.e., indirectly versus directly).

The values of **ℛ**_*E*_ for both strategies are below the corresponding **ℛ**_0_ values (figures 8a and c), of which our estimates are 5.2 and 7.7 during 2020-21 and 2022-23, respectively. As heterogeneity, especially when mixing is preferential (table 1), increases **ℛ**_0_ (Feng, et al. 2015; Hill, et al. 2023), estimates are much lower in models with fewer age classes or spatial strata and proportional mixing (table 1). While vaccination was not the only intervention modeled, all else was equal in these comparisons. As younger individuals contribute more to reproduction numbers than older ones, by virtue largely of their greater contact rates (figure 4a), the alternative reduced **ℛ**_*E*_ more than the actual strategy (figures 8b and d), approximately 19 and 8% more on average during 2020-21 and 2022-23, respectively.

## IV. Discussion

Motivated by two striking observations, we used our current model (figure 1), validated by comparing its predictions under actual conditions to accurate independent observations (figure 2), to evaluate protecting vulnerable people by vaccinating those who might otherwise infect them (i.e., indirectly).

The first observation is the gradient of **ℛ**_*E*_ with respect to vaccination before and after Q1 of 2021 (figure 3), which indicates that reducing **ℛ**_*E*_ most expeditiously requires vaccinating children. The striking resemblance of the marginal contact rates and proportions with anti-N in data collected for the October 2021 ACIP meeting (figure 3), at which vaccinating children would be considered, is second. This observation indicates that closing schools and other NPIs did not affect the contacts of schoolchildren disproportionately. Moreover, because the contact rates of older people are much lower than those of younger ones (figure 3a), even if viral load increased with symptoms, which increase with age, infected older people would contribute less to **ℛ**_*E*_ than younger ones (figures 5c and 6c), especially if hospitalized (i.e., subject to infection-control procedures).

While vaccinating younger people would therefore reduce transmission more, whether more illnesses, hospitalizations, and deaths would be averted directly or indirectly was unknown. Accordingly, we compared the impact of the reported weekly vaccinations at each location with reallocations by dose to age groups in proportion to their contributions to **ℛ**_*E*_ at the end of the previous week. We estimate that approximately 1.5×10^7^ more illnesses and 4.5×10^6^ more hospitalizations and 4.6×10^5^ more deaths were averted by the actual than alternative strategy during 2020-21 (figure 7b). The corresponding numbers during 2022-23 are lower (figure 7d) because there were fewer vaccinations during this period and a greater proportion were booster versus primary doses (figures 5a and 6a).

Thus, results of our comparison of strategies during the entire Public Health Emergency in the U.S. resemble those of our more limited earlier comparison (Vo, et al. 2023) and Bubar, et al. (2021): Given the policy goal of protecting those most vulnerable to severe disease, the actual strategy outperformed the alternative, reallocation by dose of vaccines administered at each location each week in proportion to age group contributions to the effective reproduction number the previous week.

To maintain vaccinations by dose at each location each week the same, our hypothetical strategy essentially reallocated doses from older to younger people, which would be unethical. After the initial phases, however, vaccine was not limited in the U.S., so vaccinating younger people need not have been at the expense of older ones. As vaccines were not approved for adolescents until Q2 of 2021 and children until Q4, however, this could not have begun as early as it did in our simulations.

While vaccinating younger people was less effective at protecting the older ones most vulnerable to severe COVID-19, assuming age-independent effectiveness and duration, increasing their vaccination would have other benefits. Influenza provides a precedent. Absent evidence that annual revaccination of elderly people reduced hospitalizations or deaths (Simonsen, et al., 2007), younger adults began being vaccinated in 2002, followed by adolescents and children. By 2010, influenza vaccination was universally recommended (NFID 2023), not only to reduce absenteeism in workplaces and schools, but also transmission to others. We describe the actual impact of vaccinating older children and adolescents against COVID-19 and that of hypothetically increased coverage elsewhere.

In future pandemics, vaccinating younger people could also reduce the economic costs of NPIs which, despite efforts during COVID-19 to mitigate their consequences (Skarp, et al. 2021), most affected families with lower incomes. As the infection-induced serological profile mirrors the marginal contact rates (figure 3), moreover, closing schools does not seem to have affected young people’s contacts disproportionately. It did, however, affect their educational achievement and social development (Goldhaber, et al. 2022), especially those without stable home environments (Engzell, et al. 2021).

Regulators ensure that pharmaceuticals are safe and effective via clinical trials. The greater severity of most vaccine-preventable diseases among adults alone would lead to populating trials with adults. But this also allows policymakers to ensure that vaccines are safe via post-marketing surveillance of adult recipients before conducting clinical trials among children (FDA 2020).

In future pandemics, earlier evaluation of vaccine safety among younger people might prove beneficial given their role in transmission. If teachers and administrative staff were considered essential workers, moreover, vaccinating younger people earlier might enable schools to reopen sooner, encouraging hesitant parents, especially working ones, to vaccinate their children, potentially mitigating the economic, educational, and developmental costs of school closures.

## V. Limitations

Picasso (1982) wrote, “We all know that art is not truth. Art is a lie that makes us realize truth, at least the truth that is given us to understand. The artist must know the manner whereby to convince others of the truthfulness of his lies.” Mechanistic modeling is the art of capturing the essential processes underlying complex natural phenomena in systems of equations or, to paraphrase Einstein, simplifying them as much as possible, but not more so.

As the literature rarely, if ever, contains the wherewithal to parameterize transmission models, assumptions are required. Advised by subject-matter experts (acknowledgements), we assumed that antibodies were informative about immunity, especially during the pre-Omicron era, that viral loads were informative about infectiousness, and so on. Immune senescence is well documented (Fuentes, et al. 2017; Pawelec 2018; Lee, et al. 2022; Liu, et al. 2023; Gyani, et al. 2024), but as limited available evidence suggests that the efficacy if not duration of vaccine-induced immunity to SARS-CoV-2 is age-independent, we could not assume otherwise.

We view mechanistic models as hypotheses (Glasser and Feng, 2025), so compare their predictions to accurate independent observations, and remedy the cause(s) of apparent disparities to ensure that their predictions under contrafactual conditions will be reliable. Unlike serology, however, reported cases, hospitalizations, and deaths are subject to under-if not over-reporting. Comparison of predicted and reported deaths led us to reduce mortality rates during 2022-23 by 1/3 relative to 2020-21, reflecting the availability of anti-viral drugs and otherwise improved medical care as well as pathogen evolution.

We detail our sources, assumptions, and validation of predicted infections and vaccinations in the Supplement. Nonetheless, the accuracy of our model’s contrafactual predictions may never be known.

## VI. Conclusions

Our comparison of the U.S. vaccination program, designed to mitigate disease, with one designed to mitigate infections was motivated by the possibility that vaccinating super-spreaders might compensate for the relative inefficiency of indirect protection. Absent evidence to the contrary, we assumed that vaccine effectiveness and duration of protection were age-independent, whereupon direct protection was superior. However, by virtue of the overall design of clinical trials, effectiveness in elderly and immunocompromised people will never be available when policy must be formulated. While reported age-specific durations of protection against COVID-19 are limited even now, policymakers emphasize the need for revaccination among those at risk of serious disease if infected. Why? Because there is ample evidence of immune senescence in general, if not specifically against SARS-CoV-2, and absence of evidence is not evidence of absence. Moreover, as closed schools not only burdened working parents, but deprived children of educational and social opportunities, reopening them for vaccinated students, teachers and administrative staff sooner in future would encourage parents who might otherwise be reluctant to vaccinate their children, especially working ones, with multiple concomitant benefits.

## Data Availability

All data are either published or available at the url listed.

https://covid.cdc.gov/covid-data-tracker

## Acknowledgements

The findings and conclusions in this report are those of the authors and do not necessarily represent the official positions of their respective institutions. CDC colleagues contributed ideas, ensured that our assumptions were consistent with the literature, and improved the manuscript.

Unpublished data are available at https://covid.cdc.gov/covid-data-tracker.

## Funding

We used two nationwide, CDC-funded, serological surveys. Except for Professor Day, the authors are (or were) U.S. government employees whose research is (or was) not funded independently.

## Author contributions

ZF wrote the equations, derived the analytical expressions, contributed to their programming, and revised the manuscript; TD checked her equations, improved their programming, and revised the manuscript; JWG designed the study, evaluated analytical results, programmed simulations, and wrote the manuscript. We used Mathematica, currently version 14.3, exclusively. Pending curation by Wolfram Research, our notebooks are available from the corresponding author upon request.

## Appendix

## Derivation of the Reproduction Numbers

The alternative vaccination strategy described in the main text reallocates vaccinations by dose at each location each week to the eligible people by age in proportion to the left eigenvector associated with the dominant eigenvalue (table 1) of the NGM (table 1) evaluated at the end of the previous week.

Here we derive those NGMs via the approach of Van den Driessche and Watmough (2002). To facilitate explication, the system of equations in the manuscript has a single index, representing age, location, or both, but our models actually are cross classified by age and location (i.e., the actual equations have two indices). While, consequently, we use only the reproduction numbers and related quantities for two-level mixing in this work, we begin by deriving those for one-level mixing.

### a. One-level mixing

Consider the disease vector for subgroup *i*: **{*E***_***is***_, ***E***_***ip***_, ***I***_***ia***_, ***I***_***ips***_, ***I***_***is***_, ***I***_***ih***_, ***I***_***ipm***_, ***I***_***im***_}, ***i*=1**,**2, …**,***n***. Absent disease, the susceptible classes are *S*_*i*_, *V*_*i*_, and *P*_*i*_. At the disease-free equilibrium, their densities are 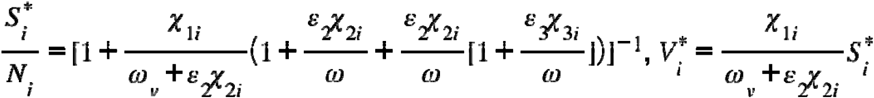, and 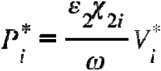.

Letting the vectors *η* be relative infectiousness, ***P*** transition probabilities and ***T*** sojourns in the several infectious states,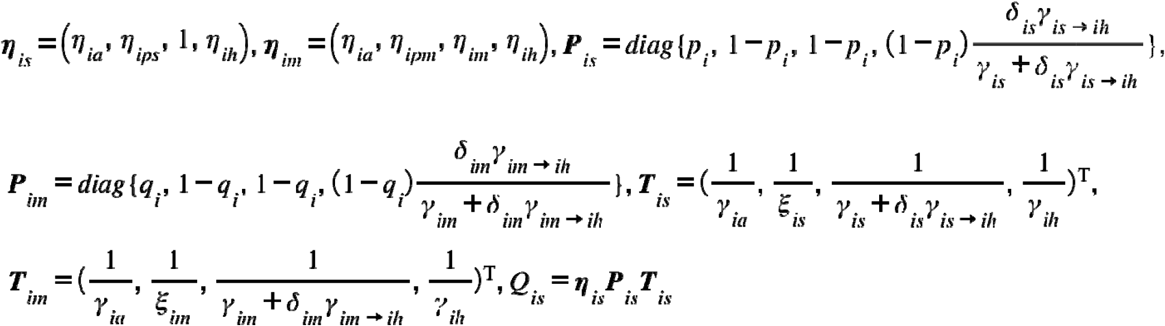 and ***Q*_*im*_= *η*_*im*_*P*_*im*_*T*_*im*_**.

If 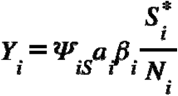 and 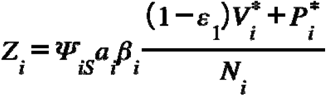 then the dominant eigenvalue of the next generation matrix (metapopulation effective reproduction number) is the same as that of

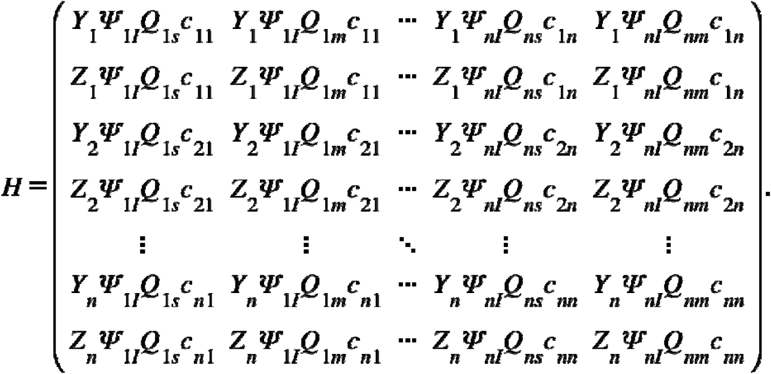

Alternatively, the matrix *H* may be written as *Ω****c****Ψ*_***I***_***Q***, where *Ω* =***diag* {*Y***_**1**_, ***Z***_**1**_, ***Y***_**2**_, ***Z***_**2**_,**…, *Y***_***n***_, ***Z***_***n***_}, ***ψ***_***I***_ **= *diag* {*ψ***_**1*I***_, ***ψ***_**1*I***_, ***ψ***_**2*I***_, ***ψ***_**2*I***,_**…, *ψ***_***nI***_, ***ψ***_***nI***_ **}, *Q* = *diag* {*Q***_**1*s***_, ***Q***_**1*m***_, ***Q***_**2*s***_, ***Q***_**2*m***_, **…, *Q***_***ns***_, ***Q***_***nm***_}, and

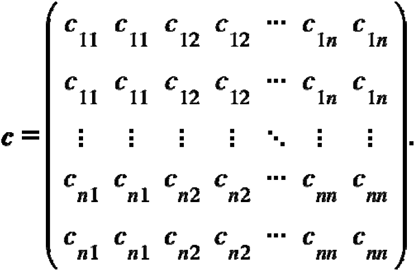

In this expression for *H*, the elements of the diagonal matrix *Ω* are rates at which fully susceptible and partially immune people are infected, ***c*** is the mixing matrix, the *Ψ*_*I*_ are reductions in infectivity by virtue of NPIs, and the ***Q*** are infectiousness-weighted sojourns. For subgroup *i*, the effective reproduction number is **ℛ**_***Ei***_ **= *Y***_***i***_ ***ψ***_***il***_ ***Q***_***is***_ **+ *Z***_***i***_ ***ψ***_***il***_ ***Q***_***im***_, ***i*= 1**,**2, …**, ***n***. For the basic number, set all *χ* = 0, *γ* = 0, and *Ψ* = 1.

### b. Two-level mixing

Consider the *m* × *n* subgroups where *m* and *n* denote numbers of locations and age-groups, respectively. In this case, the FOI for the sub-population at location *i* and age-group *j* is given by

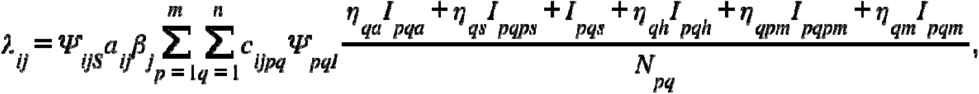

where *c*_*ijpq*_ is the proportion of the contacts of people at location *i*, age-group *j* that are with those at location *p*, age-group *q*.

Let ***v*** denote the vector of infected state variables in the following order: ***v* =( *v***_**11**_, ***v***_**12**_, **…**, ***v***_**1*n***_, ***v***_**21**_, ***v***_**22**_,**…, *v***_**2*n***_, ***v***_***m*1**_, ***v***_***m*2**_,**…, *v***_***mn***_), where ***v***_***ij***_ denotes the vector of disease stages for location *i* and age-group *j*: ***v***_***ij***_ **=( *E***_***ijs***_, ***E***_***ijP***_, ***I***_***ija***_,***I***_***ijps***_, ***I***_***ijs***_, ***I***_***ijh***_, ***I***_***ijpm***_, ***I***_***ijm***_ **), 1 ≤ *i* ≤ *m*, 1 ≤ *j* ≤ *n***. Consider the system composed of fractions of the variables within each group; e.g., *s* = *S*_*ij*_/*N*_*ij*_. In the absence of disease, the non-zero variables are given by 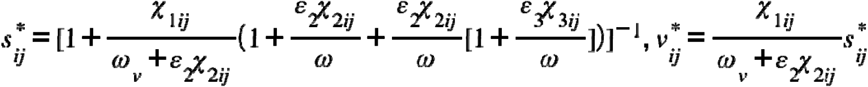, and 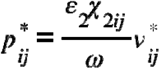.

Assume that some parameters depend on age alone (i.e., are independent of location), in which case, only a single subscript is used, e.g., ***η***_***js***_ is for age-group *j*. As for one-level mixing, letting

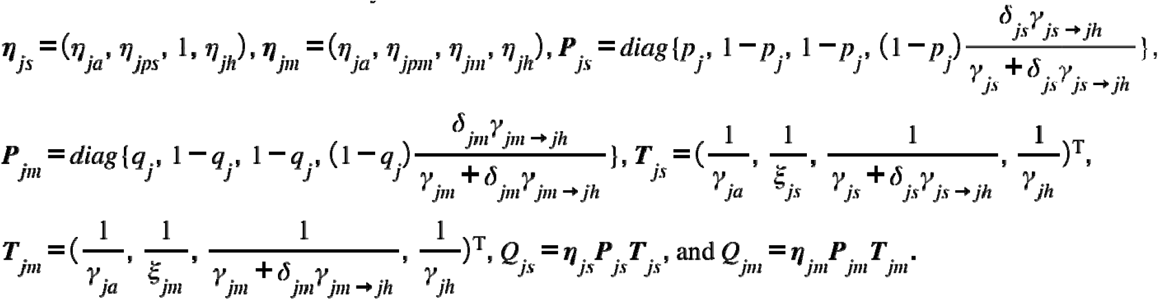

In addition, denote the quantities at location *i*, age-group *j* by

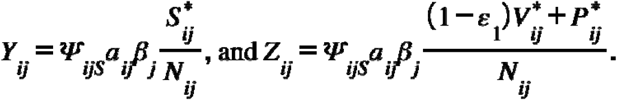

Consider the case of *n* = 3 age-groups and *m* = 2 locations. The NGM is ***FV***^**1**^ where ***F*** and ***V*** are matrices of the infection and transition terms from the Jacobian and ***F*** has the following block matrix form:

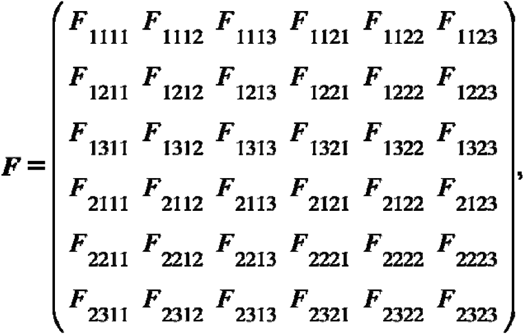

where ***F***_***ijpq***_ **(1 ≤ *i***,***p* ≤ *m* and 1 ≤ *j, q* ≤ *n*)** is an 8 × 8 matrix representing interactions between susceptible individuals at location *i*, age-group *j*, and infected individuals at location *p*, age-group *q*, given by

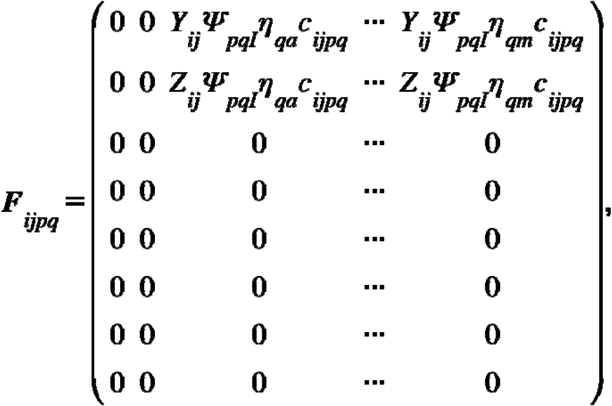

and

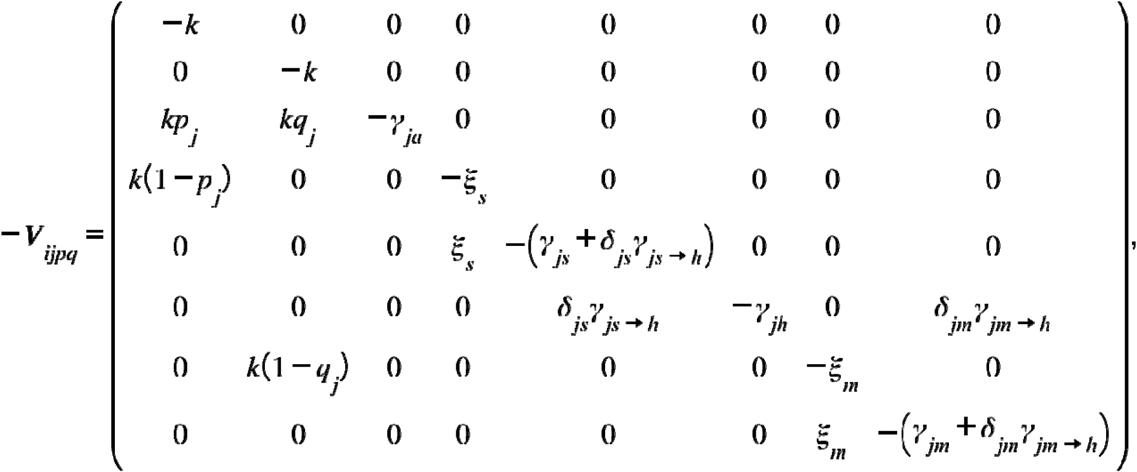

**if *i* =*p* and *j* = *q* or −*V***_***ijpq***_ **=0 if *i* ≠ *p* or *j* ≠ *q***.

For general *m* and *n*, the dominant eigenvalue of the NGM is the same as that of the following matrix:

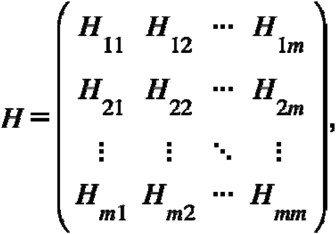

where ***H***_***ip***_ **(1 ≤ *i, p* ≤ *m*)** is a 2*n*-by-2n matrix representing interactions between susceptible individuals at location *i* and infected individuals at location *p*, given by

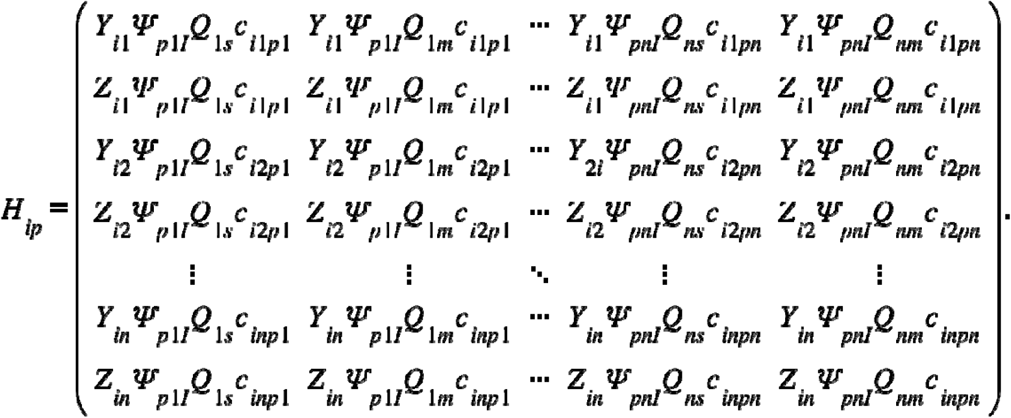

Alternatively, the matrix *H*_*ip*_ can be written as: *H*_*ip*_ = *Ω*_*i*_*c*_*ip*_*Ψ*_*pl*_*Q*_*p*_, where

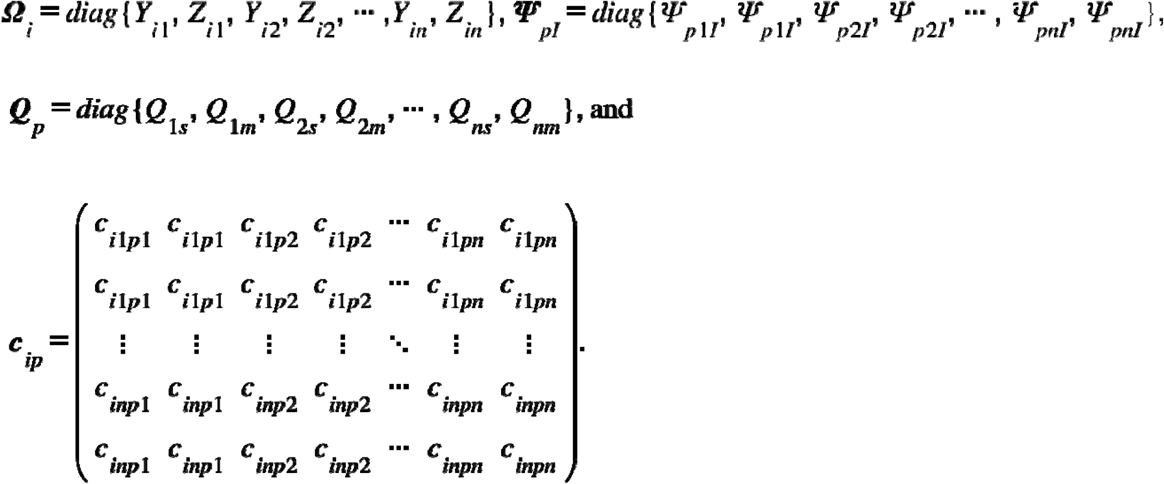

The elements of this expression have the same biological interpretations as those for one-level mixing. And the metapopulation effective reproduction number (table 1), **ℛ**_***E***_ is given by the dominant eigenvalue of *H*. Setting the parameters by which interventions are represented to zero or one as before, the metapopulation basic reproduction number, **ℛ**_**0**_ is obtained. Both associated eigenvectors have biological interpretations (Hurford, et al. 2010).

## Notes

### Competing Interest Statement

The authors have declared no competing interest.

### Summary of Updates

Revised in response to reviews.

