## Supplement for "A comparison of alternative vaccination strategies for protecting those vulnerable to illness, hospitalization, or death upon infection with SARS-CoV-2"

Introduction

During the COVID-19 pandemic, we endeavored to keep pace with understanding of biological phenomena that might affect SARS-CoV-2 transmission by modifying SEIR metapopulation models structured by age, location, or both. Using independent estimates of all but a single parameter, our age- and location-stratified models reproduce seroprevalence from a nationwide survey. Our model’s ability to reproduce those accurate independent observations (without fitting to them or any others) during 2020-21 enables us to answer reliably counter-factual questions. In the accompanying manuscript, we ask, “How would the impact of vaccinating to reduce transmission compare with the actual strategy, designed to reduce serious illnesses, hospitalizations, and deaths?”

Here we compare predictions of our current model with those accurate independent observations (Section 1), explain how its structure and parameter values differ from our published model (Section 2), and respond to questions about the utility of our approach elsewhere (Section 3). Figures and tables whose numbers are preceded by S are here; others are in the main manuscript.

Section 1. Validation

We estimated the age-specific forces of infection (table 1), attack rates (table 1), and – together with contact rates from Prem, et al. (2017) – probabilities of infection on contact (table 1) and initialized our penultimate model during Q4 of 2020 from a survey of commercial laboratory client sera (Bajema, et al. 2021). We set
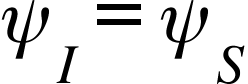
 and chose a value with which predicted seroprevalence resembled that observed just before vaccination began on 14 December 2020 (Glasser, et al. (2023). All other parameter values were informed by the literature (tables S1-S4). Judging from a survey of blood-donor sera (Jones, et al. 2021), predictions became increasingly accurate during 2021 (figure 2).

While those surveys were discontinued or repurposed at the end of 2021, we matched the timing of peak infections associated with successive Omicron variants either by one-time adjustments or temporal variation in several parameters during 2022-23, notably the duration of immunity, susceptibility to infection by successive variants, vaccine effectiveness, and compliance with NPIs (Section 2). We also compared predictions to quarterly estimates of seroprevalence (Wiegand, et al. 2023), which diverged during 2022-23 as the protection against infection with successive Omicron variants conferred by antibodies from prior infection (Chemaitelly, et al. 2025) or vaccination declined.

Section 2. Differences between our current and published SARS-CoV-2 transmission models

a. Contacts within locations

After presentations at the Spring 2023 Southeastern Sectional Meeting of the American Mathematical Society, Claus Kadelka (<https://faculty.sites.iastate.edu/ckadelka/>) suggested making the *m* state densities population weighted averages of county or county-like jurisdiction densities (similar to U.S. density, which was a population-weighted average of state densities), to better account for rural-urban heterogeneity. Consequently, figure S1 differs from Glasser, et al. (2023) figure 5. Otherwise, mixing is the same in this and our published models.

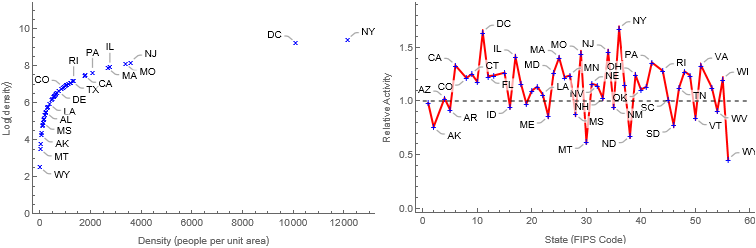

Figure S1. Multiplying contacts within locations by ratios of natural logs of population density, a) saturating functions of density, and that of the entire country, b) reduces the difference in contact rates between most and least densely populated locations to twofold.

b. Biological parameters

Except as described below, symbol definitions, values, and sources are unchanged from Glasser, et al. (2023) and Vo, et al. (2023), conserving valuable journal space.

For Q1 of 2021, we used vaccine efficacy from the clinical trial of the least effective mRNA vaccine (El Sahly, et al. 2021). We modified those for 1^st^, 2^nd^, ... doses, *ϵ*_1_, *ϵ*_2_, ... during subsequent periods to account both for the mix of products used in the U.S. and dominant variants (Braeye, et al. 2023; Feiken, et al. 2023; Voght, et al. 2023; see table S4).

As several investigators had reported reduced vaccine effectiveness with age after the first dose (e.g., Collier, et al. 2021; Rosenberg, et al. 2022), but not after the second (Jergović, et al. 2022) or third (Newman, et al. 2022; Renia, et al., 2022), we assumed that vaccine effectiveness and the duration of immunity (infection- as well as vaccination-induced) were age independent.

Originally, we assumed that immunity lasts a year. The University of Washington’s COVID-19 Forecasting Team’s meta-analysis (Stein, et al. 2023) indicates that prior infection was 78·6% protective against reinfection at 40 weeks pre-Omicron. The WHO group’s (Bobrovitz, et al. 2023) Omicron number is lower at 3 months (65·2%) and much lower at 12 months (24·7%).

In our model without immunity-moderated disease, the *p_i_* were age-specific proportions without co-morbidities that increase the risk of serious disease (from Clark, et al. 2020). We assumed that, upon infection, they would have few if any mild symptoms and their complements moderate to severe symptoms (table S1). We chose *q_i_* = 1 – (0.5 [1 – *p_i_*]), a 50% reduction in the age-specific proportions severely ill among those with partial immunity compared to immunologically naïve people (table S2).

The *δ_is_* are age-specific proportions with symptoms who require inpatient care (also from Clark, et al. 2020), which they receive after having been ill on average for 1/*γ_is→h_* = 2 days. We reduced these proportions by 50% among those with immunity-moderated (moderate) symptoms, who seek care after having been ill for an average of 1/*γ_im→h_* = 2 days (i.e., 1/*γ_im→h_* = 1/*γ_is→h_* ). Unless hospitalized, none die.

Similarly, symptoms last 1/*γ_s_* = 5 days with a 1/*ξ_s_* = 2-day pre-symptomatic period, mild illnesses also last 1/*γ_a_* = 7 days, and hospitalization lasts 1/*γ_h_* = 10 days, all averages. We assumed that the durations of pre- and moderate symptoms were the same, as was the duration of hospitalization. In other words, if inpatient care is needed, partial immunity from prior infection or vaccination doesn’t matter.

Relative to people with moderate to severe symptoms, we assumed that those with few if any mild ones were 0.5, pre-serious ones 1.25, and those hospitalized 0.1 (due to infection-control measures) as infectious. Similarly, we assumed that those with pre- and moderate symptoms were 1 and 0.75 times as infectious as those with severe symptoms (i.e., intermediate).

Multiplying IFRs from Levin, et al. (2020) by complements of the age-specific proportions without co-morbidities from Clark, et al. (2020) yields mortality rates with which predicted deaths resemble those reported during 2020-21. Some parameters in our penultimate model were time-varying (figure S2). Others that were constant became time-varying in response to the approval of less efficacious vaccines, dominance of novel variants, and host-population fatigue (table S4).

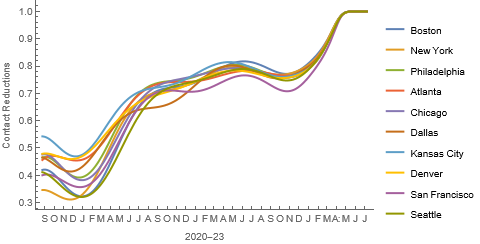

Figure S2. Complements of quotients of the Oxford Stringency Indices (Hale, et al. 2021) and 100 for HHS Regions from September 2020 through the end of the Public Health Emergency during May 2023.

Virulence may have increased as Alpha replaced Wuhan and Delta replaced Alpha (Carabelli, et al. 2023; Markov, et al. 2023), but the probabilities of hospitalization, *δ_i_* and dying, 1 – *θ_i_* were not further adjusted until Omicron replaced Delta and more efficacious treatments became available (e.g., Paxlovid during May 2023). The magnitude of this reduction (table S4) was approximated from changes in the ratios of reported hospitalizations (Devine, et al. 2024) and deaths to modeled infections.

Finally, just before the first Omicron peak, the reduced duration of protection conferred by immunity to prior variants was modeled by moving one third of those having recently Recovered, completed the primary vaccine series, state V, or been boosted to the Partially immune and one third of them to the fully Susceptible classes. This is tantamount to reducing the half-life of immunity from 6 to 4 months and is consistent with the recent report of Chemaitelly, et al. (2025).

In our reduced models (e.g., larger age-groups or census divisions or HHS regions versus states plus D.C.), parameters are population-weighted averages of those in the full model.

Section 3. In which major aspects is this model specific to the U.S. population and how could it be applied to other countries?

We began modeling SARS-CoV-2 transmission when the senior author was a member of the CDC pandemic response’s International Task Force. When the response ended, he joined a division created to assume the domestic response activities that continued (and be responsible for other viral respiratory diseases). Our goal changed from helping countries to use the resources available to mitigate the pandemic most effectively to learning if we could have responded more effectively at home.

In smaller, more homogeneous countries, we modeled NPIs (e.g., school closures, curfews, …) explicitly. In the United States, interventions were too heterogeneous in nature and timing for that, so we used Oxford Stringency Indices (figure S2). We learned from experiences. Our first U.S. model was a modification of one that we had used in Jamaica.

Our estimates of the forces of infection (Glasser, et al. 2023) are from a serosurvey in the United States (whose results are publicly available). We know that they don’t vary temporally (pre-vaccination), but are unsure if they vary spatially. Because they were estimated from first principles (versus by fitting our transmission model), however, others could use them (Glasser and Feng 2025).

Weekly numbers vaccinated by age, location, and dose were reported to the CDC (and are publicly available). We used them, together with numbers eligible for vaccination from model solutions at the end of the previous week, to model the weekly vaccination rates. Were such information not available elsewhere, the vaccination rates would have to be modeled differently.

Other required information (e.g., sub-population sizes, marginal contact rates and with whom, stringency indices (Hale, et al. 2021)) are available for many, if not most other countries.

As mixing in critically important in metapopulation modeling (Feng and Glasser 2019), there are numerous studies. Some are location-specific and others are adaptations to locations where studies may not have been conducted (e.g., Prem, et al. 2017). We recommend checking to ensure that results of the adaptations make sense. According to that of Mistry, et al. (2021), for example, parents in the U.S. have higher rates of contact with adolescents than younger children.

Most studies are based on face-to-face conversations. The number of unique contacts (i.e., between particular people) per day generally is limited, which however – unless duration is modeled – better suits them to studies of community than household transmission. For that purpose, we recommend studies based on periods in shared spaces (e.g., Del Valle, et al. 2007; Zagheni, et al. 2008).

Table S1. Symbol definitions, approximate values, and sources (modified from Glasser, et al. 2023)

| Symbol | Meaning | Values | Source |
| --- | --- | --- | --- |
| *p_i,_ q_i_* | Proportions of Susceptible and Partially-immune people whose symptoms are Mild (Severe, 1 – *p_i_* or Moderate, 1 *– q_i_*) upon infection; *q_i_* = 1 – (0.5 (1 – *p_i_*)) | Tables S2 and S3 | Clark, et al. 2020 |
| *κ* | *Per capita* rate of progression from Exposed, *E_s_* to Mildly, *I_a_*, pre-Moderately, *I_pm_*, or pre-Severely symptomatic, *I_ps_* (originally 1/3 to 1/5 days depending on variant) | 1/4 days | He, et al. 2020 |
| *ξ_s_, ξ_m_* | *Per capita* rates of progression from Pre-*I_ps_* or *I_pm_* to Severe, *I_s_* or Moderate, *I_m_* symptoms | 1/2 days | He, et al. 2020 |
| *γ_s→h_, γ_m→h_* | *Per capita* rates of progression from Severe, *I_s_* or Moderate, *I_m_* symptoms to Hospitalized, *I_h_* | 1/2 days |  |
| *γ_a_* | *Per capita* rate at which people with Mild symptoms recover (become no longer infectious) | 1/7 days |  |
| *γ_m_* | *Per capita* rate at which people with Moderate symptoms who are not hospitalized recover (again, become no longer infectious) | 1/5 days |  |
| *γ_s_* | *Per capita* rate at which people with Severe symptoms who are not hospitalized recover (again, become no longer infectious) | 1/5 days | He, et al. 2020 |
| *γ_h_* | *Per capita* rate at which Hospitalized people recover (NB: may well differ from the discharge rate) | 1/10 days |  |
| *θ_w_* | Proportions of Severely ill people who Recover (or Die, 1 – *θ_w_*), which may differ among those Hospitalized (*w* = *s*, *h*) | Table S2 | Levin, et al. 2020 |
| *δ_is_*, *δ_im_* | Proportions of people with Severe or Moderate symptoms who are Hospitalized (proportions with 2+ co-morbidities); *δ_im_* = 0.5 × *δ_is_* | Table S2 | Clark, et al. 2020 |
| *χ*_1_*_i_*, *χ*_2_*_i_*, *χ*_3_*_i_*, | *Per capita* immunization rates, 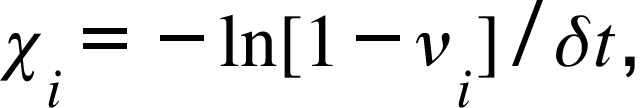where *v_i_* is the coverage attained during period *δt* | ‡ |  |
| *ϵ*_1_*, ϵ*_2_*, ϵ*_3_ | Probabilities of protection upon contact with infectious people 14 or more days after first, second, and booster doses (may be age-specific) | 0.93 | El Sahly, et al. 2021 |
| *ω, ω_v_* | *Per capita* rates at which infection- and vaccine-induced immunity is lost | 1/365 days |  |
| *c_i_* | Proportion of people complying with social distancing or mask wearing recommendations (compliance) | Table S2 | Jones 2020 |
| *b_S_*, *b_I_* | Reduced susceptibility or infectivity (effectiveness) by virtue of complying with physical-distancing where possible and mask-wearing otherwise | 0.31 | Estimated |
| *η_w_* | Scaling constants (*w* = *a*, *p_s_*, *p_m_*, *m*, *h*) representing the infectivity of the infectious states, *I_ia_*, *I_ips_*, *I_ipm_*, *I_im_, I_ih_*, relative to *I_is_* = 1 | 0.5, 1.25, 0.75, 0.5, 0.1 |  |

Notes: While we assume that people with mild and moderate symptoms are less infectious than those with severe ones, we assume that their sojourns are the same. ‡Calculated from weekly reports of doses administered and numbers eligible from model simulations, both by age and location.

Table S2. Parameter values for age-stratified transmission modeling in the United States (modified from Glasser, et al. 2023)

| Age | Population numbers | Population proportions | Pr(mild\|inf), *p_i_* | Pr(severe\|inf), *δ_i_* | Pr(death\|inf), IFR | Compliance |
| --- | --- | --- | --- | --- | --- | --- |
| 0-4 | 19,676,332 | 0.059444636 | 0.981400373 | 1.0435E-05 | 0.000008 | 0.706360069 |
| 5-9 | 20,045,152 | 0.060558887 | 0.972064267 | 0.000143099 | 0.00001312163 | 0.715211185 |
| 10-14 | 21,089,487 | 0.063713953 | 0.969763501 | 0.00027027 | 0.00002400084 | 0.71952257 |
| 15-19 | 21,242,908 | 0.064177457 | 0.960291933 | 0.003637205 | 0.00004390005 | 0.720175768 |
| 20-24 | 22,258,745 | 0.067246426 | 0.941577238 | 0.006854725 | 0.0000802978 | 0.71795013 |
| 25-29 | 23,835,330 | 0.072009485 | 0.913456364 | 0.017709616 | 0.00014687309 | 0.713574937 |
| 30-34 | 23,052,479 | 0.069644395 | 0.880594618 | 0.025217824 | 0.00026864628 | 0.707771036 |
| 35-39 | 21,615,791 | 0.065303982 | 0.839642887 | 0.035166793 | 0.00049138219 | 0.701279533 |
| 40-44 | 20,294,599 | 0.061312498 | 0.775783537 | 0.037368978 | 0.0008987895 | 0.694876881 |
| 45-49 | 20,053,798 | 0.060585008 | 0.708076043 | 0.051520404 | 0.00164398026 | 0.689376822 |
| 50-54 | 20,577,807 | 0.062168104 | 0.623043155 | 0.062486388 | 0.00300701205 | 0.685620487 |
| 55-59 | 21,542,270 | 0.065081866 | 0.521907521 | 0.083247801 | 0.00550014019 | 0.684456022 |
| 60-64 | 20,669,143 | 0.062444041 | 0.421531351 | 0.090697794 | 0.01006033295 | 0.686708224 |
| 65-69 | 17,819,027 | 0.053833488 | 0.328351424 | 0.110144916 | 0.01840140307 | 0.693136453 |
| 70-74 | 14,354,863 | 0.043367819 | 0.2503706 | 0.118491978 | 0.03365809369 | 0.704376296 |
| 75+ | 22,874,916 | 0.069107955 | 0.163464241 | 0.139755917 | 0.1430030403 | 0.744380002 |
| Wtd Avg |  |  | 0.715196 | 0.047154 | 0.0139369 | 0.705841 |

Notes: Populations, proportions without co-morbidities (assumed to have mild if any symptoms) and requiring hospitalization if infected are from Clark, et al. (2020), the IFRs are from Levin, et al. (2020), and the compliances are our estimates, obtained from Jones (2020). We have since included the decline reported by Crane, et al. (2021) and subsequent ones (table S4).

Table S3. Additional parameters associated with immunity-moderated disease.

|  | Symbol | Meaning | Values | Source |
| --- | --- | --- | --- | --- |
|  | *p_i,_ q_i_* | Proportions of Susceptible and Partially-immune people who have Mild if any symptoms (Severe, 1 – *p_i_* or Moderate symptoms, 1 *– q_i_*) upon infection; *q_i_* = 1 – (0.5 (1 – *p_i_*)) | Table S2 | Clark, et al. 2020 |
|  | *ξ_s_, ξ_m_* | *Per capita* rates of progression from Pre-*I_ps_* or *I_pm_* to Severe, *I_s_* or Moderate, *I_m_* symptoms | 1/2 days | He, et al. 2020 |
|  | *γ_s→h_,*  *γ_m→h_* | *Per capita* rates of progression from Severe, *I_s_* or Moderate, *I_m_* symptoms to Hospitalized, *I_h_* | 1/2 days |  |
|  | *γ_m_* | *Per capita* rate at which people with Moderate symptoms who are not hospitalized recover (become no longer infectious) | 1/5 days | He, et al. 2020 |
| *δ_is_*, *δ_im_* | | Proportions of people with Severe or Moderate symptoms who are Hospitalized (proportions with 2+ co-morbidities); *δ_im_* = 0.5 × *δ_is_* | Table S2 | Clark, et al. 2020 |

Table S4. Time-varying parameters (*β*, *c_i_*, *δ_i_*, and 1 – *θ_i_*, relative to Wuhan). Except as described above, symbol definitions, values, and sources are unchanged from Glasser, et al. (2023).

| Symbol | Meaning | Wuhan | Alpha | Delta | BS1 | BA5 | Source |
| --- | --- | --- | --- | --- | --- | --- | --- |
| 1/*κ* | Latent period | 5 days |  | 4 days |  | 3 days | He, et al. 2020 |
| *β_i_* | Probabilities of infection on contact with an infectious person | Fig 4 (of the source) | 1.25× | 1.5× | 2× | 1× | Glasser, et al. 2023 |
| *ϵ*_1_*, ϵ*_2_*, ϵ*_3_ | Probabilities of protection upon contact with infectious people 14 or more days after first, second, and booster doses | 0.9 | 0.8 | 0.7 | 0.6 | 0.5 | Feiken, et al. 2023 |
| *c_i_* | Proportion complying with NPI recommendations | Table S2 | 0.8× | 0.6× | 0.4× | 0.2× | Crane, et al. 2021 |
| *δ_i_*, 1 – *θ_i_* | Probabilities of hospitalization and dying | Table S2 | 1× | 1× | 0.67× | 0.67× | Carabelli, et al. 2023, Markov, et al. 2023 |

Notes: The Wuhan values (table S1, modified from Glasser, et al. 2023) changed left to right as novel variants dominated, new vaccines and medications became available, and compliance with recommended physical distancing when possible and masking otherwise declined.
